# Walking Pace, Sport Genes, and the Lung Cancer

**DOI:** 10.1101/2023.10.02.23296383

**Authors:** Mengying Wang, Xiangqi Meng, Weiliang Tian, Ruinan Sun, Siyue Wang, Yilei Qin, Janice M. Ranson, Hexiang Peng, Valerio Napolioni, Patrick W. C. Lau, Tao Wu, Jie Huang

**Author notes:** Correspondence to: Jie Huang, School of Public Health and Emergency Management, Southern University of Science and Technology, Shenzhen, China; Tao Wu, Department of Epidemiology and Biostatistics, School of Public Health, Peking University, 38 Xueyuan Road, Haidian District, Beijing, China.

## Abstract

**Background:** To investigate the association between walking pace and lung cancer risk, and explore whether any association is modified by endurance and power-related genes.

**Methods:** We followed up 449,890 UK Biobank participants free of cancer at baseline. Data on self-reported walking pace were collected by touchscreen questionnaire at baseline. Blood samples were obtained for genotyping. Hazard ratios (HRs) and 95% confidence intervals (CIs) were calculated for lung cancer incidence and mortality, with slow walking pace as the reference.

**Findings:** 4,087 lung cancer incident cases and 2,245 lung cancer deaths were identified during a median follow-up period of 12.1 and 12.7 years, respectively. For incident lung cancer, HR (95% CI) were 0.71 (0.65- 0.78) and 0.55 (0.49-0.61) among participants with steady and brisk walking pace, respectively. For lung cancer mortality, steady and brisk walking paces were associated with 32% and 48% lower risks, respectively. Associations of walking pace with risks of lung cancer occurrence and mortality were modified by rs1815739 in *ACTN3* and rs7191721 in *RBFOX1*. The protective effect of faster walking pace was more evident among participants carrying a higher number of T allele for rs1815739 in *ACTN3* (*P*_interaction_=0.04 for both lung cancer incidence and mortality) and A allele for rs7191721 in *RBFOX1* (*P*_interaction_=0.01 for lung cancer incidence, *P*_interaction_=0.004 for lung cancer mortality).

**Interpretation:** Faster walking pace is associated with lower risks of both lung cancer occurrence and mortality, and this protective effect is modulated by polymorphisms in endurance gene *RBFOX1* and in power gene (*ACTN3*).

**Funding:** This work was supported by grants from the China Postdoctoral Science Foundation (Grant No. BX2021021, 2022M710249), Fujian Provincial Health Technology Project (Grant No. 2020CXB009), and the Natural Science Foundation of Fujian Province, China (Grant No. 2021J01352).

## Introduction

Lung cancer is the leading cause of cancer incidence and mortality worldwide.^1^ Previous studies have shown that adopting healthy lifestyle behaviors is crucial for the primary prevention and reduction of mortality from lung cancer.^2–4^ In particular, physical activity has been recognized as a modifiable lifestyle factor that affects both the incidence and prognosis of the disease.^5,6^ The frequency and intensity of physical activity can be attributed to physical fitness, which consists of cardiorespiratory fitness and muscle strength.^7^ Therefore, physical fitness is reported to be an important risk factor for both the occurrence and mortality of lung cancer.^8,9^

Walking pace is a key indicator of physical fitness. Epidemiological and genetic studies have suggested that walking pace is associated with cardiovascular disease,^7,10^ type 2 diabetes,^11^ and all-cause mortality.^12^ However, the association between habitual walking pace and the risk of lung cancer incidence and mortality has been less frequently and inconsistently reported.^8,13^ In addition, previous studies have demonstrated that the relationship between walking pace and cancer risk is likely to be modified by sociodemographic factors and lifestyle choices.^7,12,14,15^ For example, Stamatakis et al. found that the association between walking pace and all-cause mortality was modified by age and physical activity.^12^

Genetic susceptibility also appears to interact with behavioral factors in relation to the risk of developing lung cancer.^14,15^ Notably, multiple endurance- and power-related genes have been identified in genome-wide association studies (GWAS).^16^ For instance, *RBFOX1* is reported to be associated with muscular dystrophies.^17^ Furthermore, the *R577X* variant (rs1815739) in the α-actinin-3 (*ACTN3*) gene is a well-recognized genetic biomarker in exercise studies.^18,19^ The C to T base substitution in *ACTN3* leads to the transformation of an arginine base (R) to a premature stop codon (X) at amino acid position 577 (*R577X*).^20^ XX individuals with the TT genotype completely lack the expression of *ACTN3,* which may influence the presence of fast fibers in the body.^20,21^ The polymorphism of the gene has been related to speed and cardiometabolic fitness.^19,22^ To the best of our knowledge, no study has evaluated the interaction between these genetic polymorphisms and walking speed in relation to lung cancer risk.

In the current study, we aimed to investigate the associations between walking pace and the risks of developing lung cancer and mortality in a substantial prospective cohort study. We also aim to assess the potential modifying effects of demographics, lifestyles, and endurance- and power-related genes on the association between walking pace and lung cancer risk.

## Methods

### Study design and participants

The current analysis was based on data from the UK Biobank study. The detailed study design and procedures were described previously.^23^ In brief, over 0.5 million participants aged 37 to 73 years were assessed at one of the 22 centers across England, Wales, and Scotland from 2006 to 2010. All participants completed a touchscreen questionnaire and physical measurements, as well as provided biological samples at the baseline survey. All participants provided written informed consent during the baseline visit. Ethical approval for the UK Biobank study was obtained from the National Information Governance Board for Health and Social Care and the National Health Service North West Multi-Centre Research Ethics Committee (Ref 11/NW/0382).

Among the 502,618 participants with available data, 449,890 individuals were included in the final analysis after excluding those with any type of cancer (N=48,535) or missing data on self-reported walking pace (N=4,193). For the genetic analysis, only participants with complete data on genetic polymorphisms were included (N=411,647).

### Assessment of outcomes

#### Lung cancer

Incident lung cancer cases were identified using hospital admission records linked to Health Episode Statistics in England and Wales and the Scottish Morbidity Records in Scotland. Incident lung cancer was defined as a hospital admission with an International Classification of Diseases, Tenth Revision (ICD-10) code of C34.

#### Lung cancer deaths

Causes and dates of death were identified by using the death certificates held by the National Health Service Information Center (England and Wales) and the National Health Service Central Register (Scotland). Lung cancer deaths were ascertained using the ICD-10 code C34.

### Assessment of walking pace

The information on self-reported walking pace was collected using the baseline touchscreen questionnaire. All participants were asked the question, “How would you describe your usual walking pace?” with “slow,” “steady,” and “brisk” paces as the response options. Slow pace was defined as <3 miles per hour; steady pace was defined as 3 to 4 miles per hour; and brisk pace was defined as >4 miles per hour.

### Ascertainment of covariates

All models were adjusted for various sociodemographic factors (age, sex, assessment center, ethnicity, UK Biobank assessment center, and Townsend Deprivation Index), lifestyles (smoking, alcohol consumption, diet, and total physical activity), health status (body mass index [BMI], grip strength, hypertension, diabetes, and cardiovascular disease), and family history of lung cancer.

The Townsend Deprivation Index is a comprehensive indicator of deprivation that includes measures of unemployment, overcrowded households, non-car ownership, and non-home ownership.^24^ Smoking and alcohol consumption status was self-reported as never, former, or current. Dietary factors were assessed using a previously published healthy diet score. One point was assigned for each favorable dietary factor based on the corresponding median intake of vegetables, fruits, fish, unprocessed red meat, and processed meat, with the total score ranging from 0 to 5.^25^ Physical activity was classified as low, moderate, or high, depending on the frequency and intensity of walking, moderate activity, vigorous activity, and Metabolic Equivalent Task (MET)- minutes/week, using items from the short International Physical Activity Questionnaire form. Detailed information about physical activity measurements is available online at https://biobank.ndph.ox.ac.uk/ukb/ukb/docs/ipaq_analysis.pdf. Height and body weight were measured by trained investigators during the baseline visit. BMI was calculated by dividing the weight (kg) by the square of the height (m). Grip strength was measured using a Jamar J00105 hydraulic hand dynamometer. The mean of the three measurements for each hand was calculated, and the maximum value from both hands was included in the analysis. A history of hypertension, diabetes, and cardiovascular disease was defined based on self-reported information and linked medical records at the time of the baseline visit. Information on family history was also collected from the touchscreen questionnaire. Family history of lung cancer was defined by an affirmative response to any of the following three questions: “Has/did your father ever suffer from lung cancer?”; “Has/did your mother ever suffer from lung cancer?”; and “Have any of your brothers or sisters suffered from lung cancer?”.

### Genotyping

The genotyping process of the UK Biobank study has been described elsewhere.^26^ In the present study, only 411,647 individuals of European descent with available genotyping data were included in the genetic analysis.

### Statistical analyses

In the analysis of the association between walking pace and lung cancer occurrence, the survival time for each participant was calculated as the duration from the response date of the baseline survey until the time of incident lung cancer, death, or the censoring date (7 April 2021), whichever occurred first. For lung cancer mortality, survival time was calculated as the duration from the baseline survey date until the date of death or the censoring date, whichever came first. The Cox proportional hazards regression model was adopted to calculate hazard ratios (HRs) and 95% confidence intervals (CIs) for each outcome. We divided participants into three categories based on their self-reported walking pace: slow, steady, and brisk. The slow-walking pace was used as the reference group. Several multivariable models were adopted to account for potential confounding factors. In the first model, all HRs were adjusted for sociodemographic factors, namely age (continuous), sex (male, female), race (white European, mixed, South Asian, black, others), UK Biobank assessment center, and Townsend Deprivation Index (continuous). In the second model, we further adjusted for alcohol consumption (current, former, never), smoking status (current, former, never), physical activity level (low, moderate, high), and healthy diet score (0, 1, 2, 3, 4, and 5). Next, we also adjusted for health indicators, including BMI (continuous), grip strength (continuous), hypertension (yes/no), diabetes (yes/no), cardiovascular disease (yes/no), and family history of lung cancer (yes/no). In the genetic analysis, we also adjusted for the genotyping batch and the first ten principal components of genetic ancestry. Walking pace was included as a continuous variable when testing for the linear trend. To handle missing covariates, we employed a missing indicator for categorical variables and imputed the mean values for continuous variables, respectively.

### Secondary analyses

Stratified analyses were performed a priori by treating walking pace as a continuous variable, taking into account age (<65 and ≥65 years), sex (male or female), Townsend Deprivation Index (<-2.1 or ≥-2.1), smoking status (non-current or current), alcohol consumption (non-current or current), physical activity (low, moderate, high), and BMI (18.5–24.9, 25.0–29.9, or ≥30 kg/m^2^). In particular, we conducted stratified analyses based on the genotype of endurance- and power-related genes. Tests for interaction were carried out by setting variable cross-product terms between walking pace and each category in the third multivariable model. The likelihood ratio test was adopted to compare models with and without the cross-product terms.

We also conducted several sensitivity analyses to assess the robustness of the results. These included: (1) adjusting for particulate matter with diameters ≤2.5 µm (PM_2.5_) exposure at baseline; (2) adjusting for body fat-free mass, calculated as the sum of muscle mass from four limbs divided by the squared height (kg/m^2^); (3) excluding participants with hypertension, diabetes, or cardiovascular disease at baseline; and (4) excluding events occurring in the first two years of follow-up to minimize potential reverse causality.

All analyses were conducted using SAS software (version 9.4; SAS Institute Inc., Cary, NC, USA). All *p*-values for the tests were two-sided, and *p*-values <0.05 were considered statistically significant.

### Patient and public involvement

Patients or the public were not involved in the design, conduct, reporting, or dissemination plans of our research.

## Results

Table 1 presents the baseline characteristics of the participants in the study. In total, 449,890 participants were included in the primary analysis, among whom 7.9%, 52.7%, and 39.4% had slow, steady, and brisk walking paces, respectively. The mean age was 56.2 (SD=8.1 years), and 240,135 (53.4%) were females. Participants with a faster walking pace had a lower Townsend Deprivation Index and were less likely to be current smokers. However, they were more likely to consume alcohol, eat healthy foods, and engage in regular exercise.

**Table 1.** Baseline characteristics of participants according to walking pace in the UK Biobank study.

| Characteristics | Walking pace |  |  |
| --- | --- | --- | --- |
|  | Slow | Steady | Brisk |
| N | 35 450 | 237 271 | 177 169 |
| Age (years) | 58.5 | 56.8 | 54.9 |
| Sex, male (%) | 45.6 | 46.4 | 47.1 |
| Race, white (%) | 88.3 | 93.4 | 96.2 |
| Townsend Deprivation Index | 0.1 | -1.3 | -1.6 |
| Current smokers (%) | 17.7 | 10.9 | 8.8 |
| Current drinkers (%) | 81.3 | 91.9 | 94.3 |
| Physical activity level (%) |  |  |  |
| Low | 31.2 | 15.5 | 11.1 |
| Moderate | 25.7 | 33.1 | 34.0 |
| High | 14.0 | 30.2 | 39.7 |
| Healthy diet score |  |  |  |
| 0-1 | 16.1 | 13.2 | 9.9 |
| 2-3 | 52.7 | 50.7 | 46.0 |
| 4-5 | 31.3 | 36.2 | 44.1 |
| Body mass index (kg/m<sup>2</sup>) | 31.2 | 28.0 | 25.9 |
| Hand grip strength (kg) | 28.3 | 32.5 | 34.4 |
| Hypertension history (%) | 68.9 | 57.0 | 46.8 |
| Diabetes history (%) | 15.3 | 5.6 | 2.6 |
| Cardiovascular diseases history (%) | 20.7 | 7.0 | 3.8 |
| Family history of lung cancer (%) | 14.8 | 12.6 | 11.2 |

Additionally, participants with a faster walking pace had a lower BMI and stronger hand grip strength. Hypertension, diabetes, cardiovascular disease, and a family history of lung cancer were less prevalent among participants with a faster walking pace.

During a median follow-up period of 12.1 years (5,349,719 person-years), 4,087 incident cases of lung cancer were identified. Additionally, during a median follow-up period of 12.7 years (5,586,963 person-years), 2,245 deaths related to lung cancer were identified.

The association between walking pace and the occurrence of lung cancer is shown in Table 2. In total, 765, 2331, and 991 incident cases of lung cancer were identified in the slow, steady, and brisk walking pace groups, respectively. We found that a faster walking pace was associated with a lower risk of developing lung cancer in all multivariable-adjusted models. In model 1, adjusted for age, sex, race, UK Biobank Assessment Centre, and Townsend Deprivation Index, the HR (95% CI) was 0.56 (0.52–0.61) for individuals with a steady walking pace and 0.39 (0.35–0.42) for individuals with a brisk walking pace compared to those with a slow walking pace (*P* trend <0.001). After further adjustment for alcohol consumption, smoking status, physical activity, and the healthy diet score, compared with slow walking pace, participants reported steady and brisk walking pace had HRs of 0.70 (0.64–0.76) and 0.55 (0.50–0.61), respectively (*P* trend <0.001). In model 3, after adjusting for BMI, hand grip strength, hypertension, diabetes, cardiovascular diseases, and family history of lung cancer, the corresponding HRs were 0.71 (0.65–0.78) and 0.55 (0.49–0.61) for participants with a steady and brisk walking pace, respectively (*P* trend <0.001).

**Table 2.** Adjusted HRs and 95% CI for walking pace with the risk of incident lung cancer in the UK Biobank study.

|  | Walking pace |  |  | *P* for trend |
| --- | --- | --- | --- | --- |
|  | Slow | Steady | Brisk |  |
| Cases/N | 765/35 450 | 2331/237 271 | 991/177 169 |  |
| Model 1 <sup>a</sup> | 1.00 | 0.56 (0.52-0.61) | 0.39 (0.35-0.42) | <0.001 |
| Model 2 <sup>b</sup> | 1.00 | 0.70 (0.64-0.76) | 0.55 (0.50-0.61) | <0.001 |
| Model 3 <sup>c</sup> | 1.00 | 0.71 (0.65-0.78) | 0.55 (0.49-0.61) | <0.001 |

<sup>a</sup> Adjusted for age (continuous, years), sex (male, female), race (white European, mixed, South Asian, black, others), UK Biobank assessment centre, and Townsend Deprivation Index (continuous).
<sup>b</sup> Adjusted for age (continuous, years), sex (male, female), race (white European, mixed, South Asian, black, others), UK Biobank assessment centre, Townsend Deprivation Index (continuous), alcohol consumption (current, former, never), smoking status (current, former, never), physical activity (low, moderate, high), and healthy diet score (0, 1, 2, 3, 4, 5).
<sup>c</sup> Adjusted for age (continuous, years), sex (male, female), race (white European, mixed, South Asian, black, others), UK Biobank assessment centre, Townsend Deprivation Index (continuous), alcohol consumption (current, former, never), smoking status (current, former, never), physical activity (low, moderate, high), healthy diet score (0, 1, 2, 3, 4, 5), body mass index (continuous, kg/m<sup>2</sup>), Hand grip strength (continuous, kg), baseline hypertension (yes/no), baseline diabetes (yes/no), baseline cardiovascular diseases (yes/no), and family history of lung cancer (yes/no).

A similar association pattern was found between walking pace and lung cancer mortality (Table 3). In total, 498, 1390, and 567 deaths from lung cancer were identified in the groups with slow, steady, and brisk walking paces, respectively. In model 1, we observed a 48% and 66% lower risk of lung cancer mortality in groups with a steady and brisk walking pace, respectively, compared to those with a slow walking pace (*P* trend <0.001). In model 3, adjusted for age, sex, race, UK Biobank Assessment Centre, Townsend Deprivation Index, alcohol consumption, smoking status, physical activity, healthy diet score, BMI, hand grip strength, hypertension, diabetes, cardiovascular diseases, and family history of lung cancer, the lung cancer mortality rate was reduced by 32% and 48% for those with a steady and brisk walking pace, respectively, when compared with a slow walking pace (*P* trend <0.001).

**Table 3.** Adjusted HRs and 95% CI for walking pace with the lung cancer mortality in the UK.

|  | Walking pace |  |  |  |
| --- | --- | --- | --- | --- |
|  | Slow | Steady | Brisk | *P* for trend |
| Cases/N | 498/35 450 | 1390/237 271 | 567/177 169 |  |
| Model 1 <sup>a</sup> | 1.00 | 0.52 (0.47-0.58) | 0.34 (0.30-0.39) | <0.001 |
| Model 2 <sup>b</sup> | 1.00 | 0.68 (0.61-0.76) | 0.52 (0.45-0.59) | <0.001 |
| Model 3 <sup>c</sup> | 1.00 | 0.68 (0.61-0.76) | 0.52 (0.45-0.59) | <0.001 |

<sup>a</sup> Adjusted for age (continuous, years), sex (male, female), race (white European, mixed, South Asian, black, others), UK Biobank assessment centre, and Townsend Deprivation Index (continuous).
<sup>b</sup> Adjusted for age (continuous, years), sex (male, female), race (white European, mixed, South Asian, black, others), UK Biobank assessment centre, Townsend Deprivation Index (continuous), alcohol consumption (current, former, never), smoking status (current, former, never), physical activity (low, moderate, high), and healthy diet score (0, 1, 2, 3, 4, 5).
<sup>c</sup> Adjusted for age (continuous, years), sex (male, female), race (white European, mixed, South Asian, black, others), UK Biobank assessment centre, Townsend Deprivation Index (continuous), alcohol consumption (current, former, never), smoking status (current, former, never), physical activity (low, moderate, high), healthy diet score (0, 1, 2, 3, 4, 5), body mass index (continuous, kg/m<sup>2</sup>), Hand grip strength (continuous, kg), baseline hypertension (yes/no), baseline diabetes (yes/no), baseline cardiovascular diseases (yes/no), and family history of lung cancer (yes/no).

We examined the interactions between walking pace and various sociodemographic and lifestyle factors. The associations between walking pace and lung cancer occurrence and mortality remained consistent across subgroups of participants, stratified by sex, age, Townsend Deprivation Index, smoking, alcohol consumption, physical activity, and BMI (Supplemental Figure 1 and Figure 2).

We further explored the interaction between walking pace and endurance- and power-related genes and the risk of lung cancer. We found that the associations between walking pace and the risks of lung cancer occurrence and mortality were modified by rs1815739 in *ACTN3* and rs7191721 in *RBFOX1.* For rs1815739 polymorphisms, the protective HR of faster walking pace was more evident among participants carrying a higher number of T alleles (*P*_interaction_=0.04 for both lung cancer incidence and mortality). The stratified analysis according to rs1815739 genotype showed that the HRs (95% CIs) for incident lung cancer associated with steady and brisk walking pace were as follows: 0.89 (0.75–1.06) and 0.65 (0.53–0.80) among participants with CC genotype; 0.67 (0.59–0.77) and 0.53 (0.46–0.63) among participants with CT genotype; and 0.60 (0.49– 0.73) and 0.47 (0.36–0.60) among participants with TT genotype, respectively (Figure 1). Similar interaction patterns were observed in the analysis of lung cancer mortality. Within the brisk walking pace group, the mortality rate for lung cancer was reduced by 41%, 48%, and 57% for carriers of the CC, CT, and TT genotypes, respectively (Supplementary Figure 3). For rs7191721 polymorphisms, the protective HR associated with a faster walking pace was stronger among participants carrying a higher number of A alleles (*P*_interaction_=0.01 for lung cancer incidence, *P*_interaction_=0.004 for lung cancer mortality). The subgroup analysis by the rs7191721 genotype showed that the HRs (95% CIs) for incident lung cancer associated with steady and brisk walking pace were as follows: 0.79 (0.66–0.95) and 0.63 (0.51–0.78) among participants with GG genotype; 0.74 (0.64–0.84) and 0.57 (0.48–0.66) among participants with GA genotype; and 0.58 (0.48–0.70) and 0.43 (0.34–0.54) among participants with AA genotype, respectively (Figure 2). Similar interaction patterns were also observed for lung cancer mortality. In the brisk walking pace group, lung cancer mortality rates were reduced by 40%, 43%, and 65% for carriers with the GG, GA, and AA genotypes, respectively (Supplementary Figure 4).

**Figure 1.**
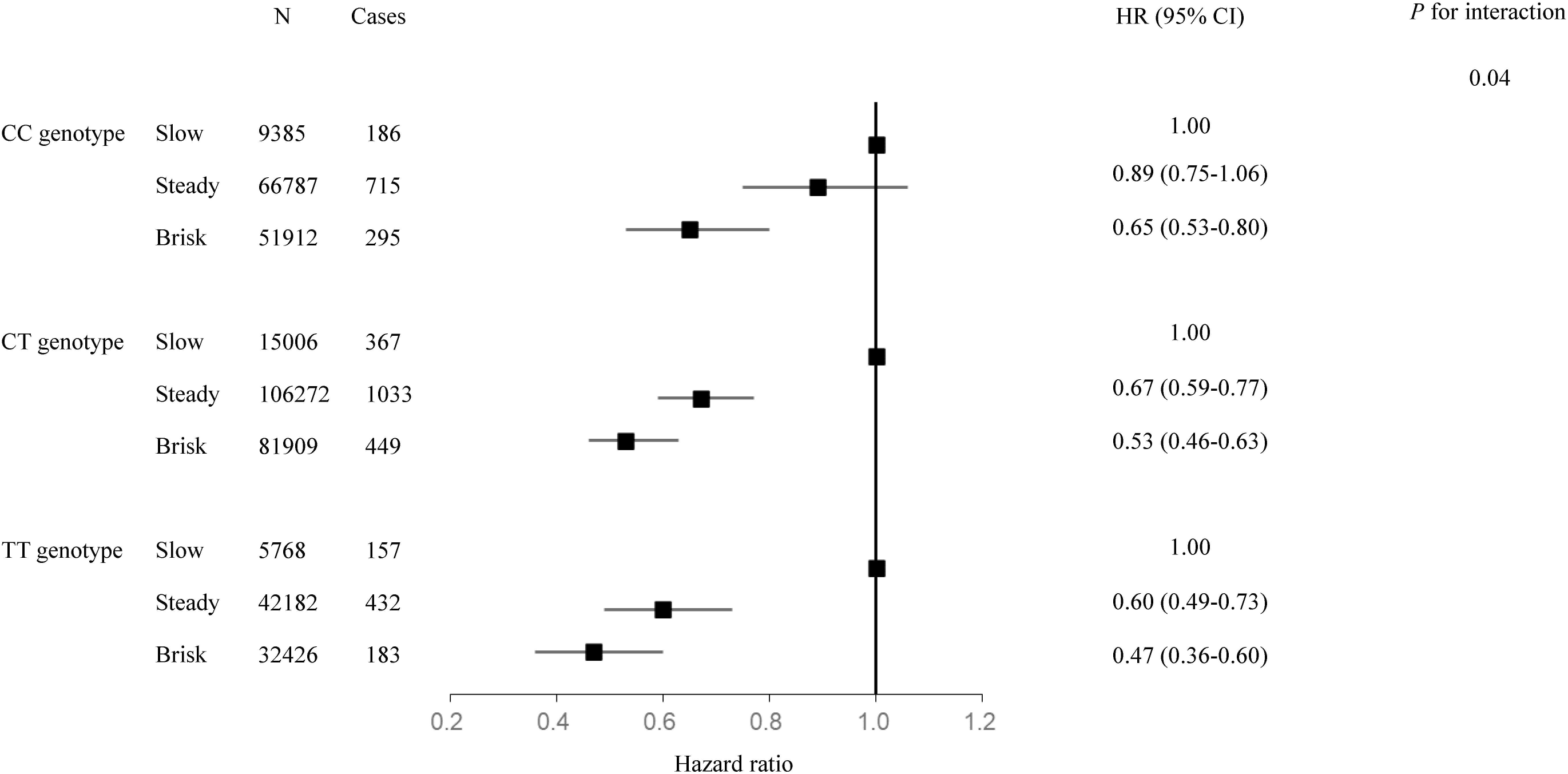
The association between walking pace and the risk of incident lung cancer stratified by rs1815739 polymorphism in *ACTN3* gene. Adjusted for age (continuous, years), sex (male, female), race (white European, mixed, South Asian, black, others), UK Biobank assessment centre, Townsend Deprivation Index (continuous), alcohol consumption (current, former, never), smoking status (current, former, never), physical activity (low, moderate, high), healthy diet score (0, 1, 2, 3, 4, 5), body mass index (continuous, kg/m^2^), Hand grip strength (continuous, kg), baseline hypertension (yes/no), baseline diabetes (yes/no), baseline cardiovascular diseases (yes/no), and family history of lung cancer (yes/no).

**Figure 2.**
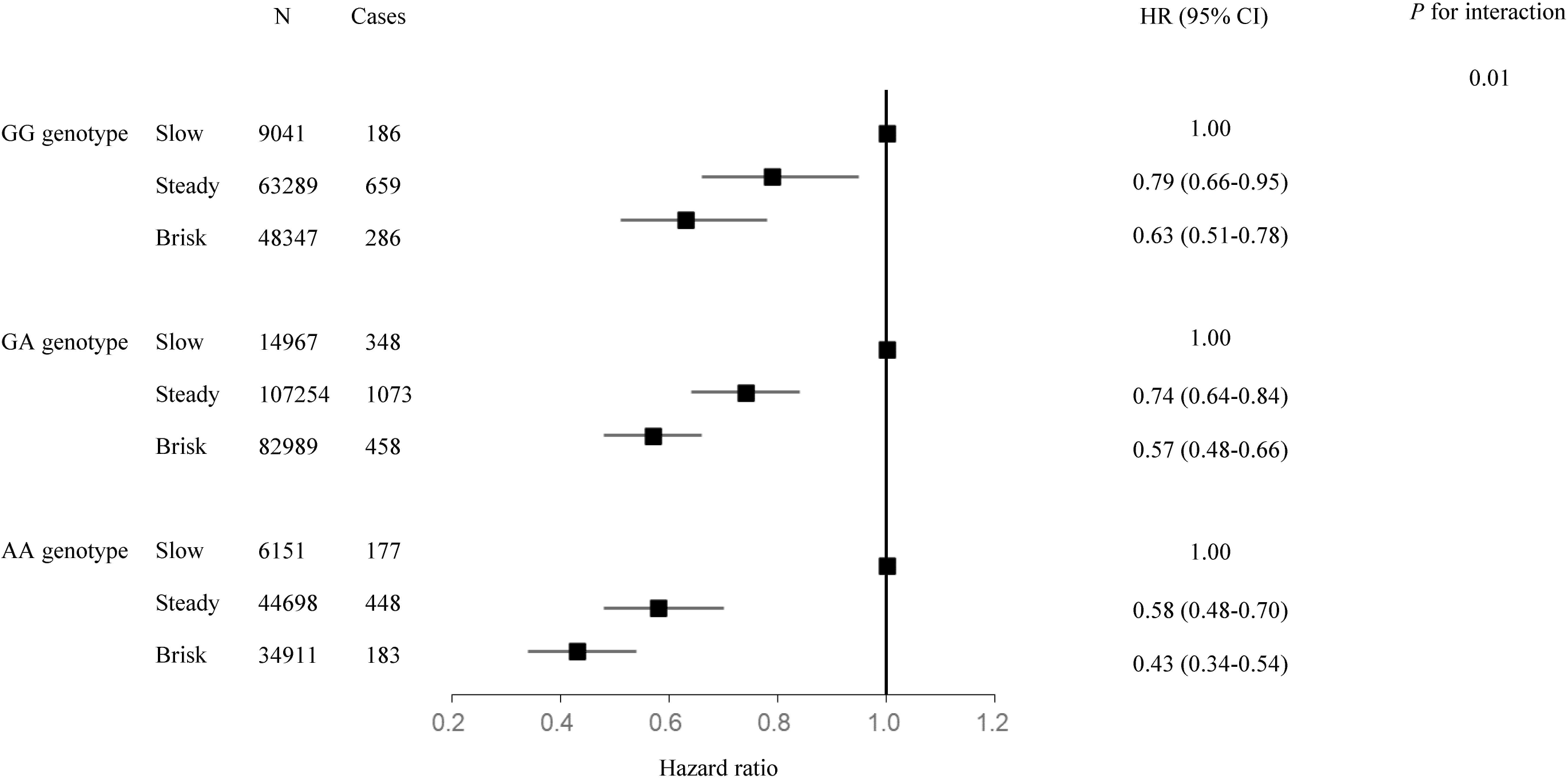
The association between walking pace and the risk of incident lung cancer stratified by rs7191721 polymorphism in *RBFOX1* gene. Adjusted for age (continuous, years), sex (male, female), race (white European, mixed, South Asian, black, others), UK Biobank assessment centre, Townsend Deprivation Index (continuous), alcohol consumption (current, former, never), smoking status (current, former, never), physical activity (low, moderate, high), healthy diet score (0, 1, 2, 3, 4, 5), body mass index (continuous, kg/m^2^), Hand grip strength (continuous, kg), baseline hypertension (yes/no), baseline diabetes (yes/no), baseline cardiovascular diseases (yes/no), and family history of lung cancer (yes/no).

The sensitivity analyses showed that additional adjustments for PM_2.5_ may not significantly change the results (Supplementary Table 1). Additionally, the associations remained significant even after adjusting for fat-free mass (Supplementary Table 2). Furthermore, after excluding participants who had hypertension, diabetes, or cardiovascular disease at baseline, the results remained stable (Supplementary Table 3). Moreover, the results remained largely unchanged when only participants with a follow-up time of more than two years were included (Supplementary Table 4).

## Discussion

In this prospective cohort study, we observed that a faster walking pace was associated with lower risks of lung cancer occurrence and mortality, independent of lifestyle factors, BMI, and hand grip strength. Of note, we found interactions between walking pace and rs1815739 in *ACTN3* as well as rs7191721 in *RBFOX1* in relation to incident lung cancer and disease mortality. Specifically, we observed that the lower risk of lung cancer associated with a faster walking pace was more prominent in individuals who carried the T allele in rs1815739 and the A allele in rs7191721, respectively. However, the associations were not influenced by sociodemographic or lifestyle factors.

The associations between walking pace and the occurrence and mortality of lung cancer have been documented in several epidemiological studies, as well as in meta-analyses of prospective cohorts, yet conflicting results have been reported.^8,13,27,28^ Similar to our findings, Smith et al. found that a slower walking pace was associated with a higher risk of lung cancer mortality compared to a faster walking pace.^27^ In addition, the results from the 40-year follow-up of the Whitehall study also indicated a significant linear trend. A faster walking pace was found to be associated with a lower risk of lung cancer mortality compared to a slower walking pace.^28^ However, a recent meta-analysis summarizing the evidence from available cohort studies on walking pace and lung cancer mortality found that slower walking speeds were not associated with lung cancer mortality.^8^ Notably, a previous analysis conducted in the UK Biobank did not find any associations between walking pace and the occurrence and mortality of lung cancer. Considering the limited number of cases and shorter follow-up time in the previous analysis,^7^ the current analysis may yield more reliable results regarding the association between walking pace and lung cancer risk.

The reasons for the observed inverse association between a faster walking pace and reduced risks of lung cancer incidence and mortality remain unknown. As suggested by a meta-analysis of randomized controlled trials, a brisk walking pace might enhance aerobic fitness,^29^ thus improving respiratory health. A previous study also demonstrates a strong dose-response relationship between walking pace and maximal oxygen uptake.^7^ These data indicate that walking pace is considered an important marker of cardiorespiratory fitness, which has been shown to be linked to a lower incidence and mortality rate of lung cancer.^8^ Additionally, exercise, including walking, might reduce the risk of lung cancer by preventing the inflammatory process.^30^

Walking is the most popular physical activity, accepted and accessible to almost the entire population.^31^ In addition, among individuals with adequate physical capacity, the change in walking pace is more practical than an increase in walking time. Therefore, the study has important public health implications for physical activity recommendations in lung cancer prevention and control. It suggests that guidelines should encourage people to walk at a faster pace in order to obtain more benefits in preventing lung cancer and reducing the risk of mortality.^32^

We did not find evidence of interaction based on sex, age, Townsend Deprivation Index, smoking status, alcohol consumption, physical activity, or BMI in the stratified analyses. Interestingly, we observed significant interactions between walking pace and two genetic variants, rs1815739 in *ACTN3* and rs7191721 in *RBFOX1*, in relation to lung cancer occurrence and mortality. Specifically, we found that the inverse association between walking pace and lung cancer risk was stronger in participants who carried the T allele in rs1815739 and the A allele in rs7191721. The *ACTN3* gene rs1815739 polymorphism occurs in approximately 16% of the global population.^20,33^ The muscle proteins α-actinins 1-4 play a crucial role in regulating the integrity of sarcomeres by binding to actin filaments in skeletal muscle. Furthermore, α-actinin proteins mediate the binding of several glycolytic enzymes to actin filaments.^34,35^ α-actinin-3, encoded by the *ACTN3* gene, is only expressed in fast fibers.^20,21^ Several epidemiological and experimental studies have suggested that the rs1815739 polymorphism in *ACTN3* is related to sports performance, especially speed.^18,19,22^ *ACTN3* has been recognized as a “speed gene,” which is associated with both endurance and power.^16,33,36,37^ In addition, the presence of rs7191721 in the *RBFOX1* gene serves as a marker for aerobic performance and endurance athlete status.^16,38^ The RNA-binding protein fox-1 homolog (C. elegans) 1 (RBFOX1), encoded by the *RBFOX1* gene, plays a key role in muscle tissue development.^39^ RNA-binding proteins are important regulators of gene expression, and Rbfox1 knockdown may result in the inhibition of muscle differentiation.^39^ These data indicate that the interactions between walking pace and these genes are biologically plausible. Our results may provide insights into the inconsistent observations regarding the relationship between walking pace and lung cancer risk in previous observational studies, where genetic modifications were not considered.

The main strengths of our study include the large population-based sample and the abundance of information on sociodemographic factors, lifestyles, and other covariates. This allowed us to conduct comprehensive stratified analyses and sensitivity analyses. Additionally, for the first time, we considered the modifying effects of genetic variations related to exercise on the association between walking pace and lung cancer risk. Several potential limitations, however, warrant consideration. Although we controlled for demographic and lifestyle factors in the analyses, there may still be unknown residual confounding. The walking pace was based on self-reported information. However, it is worth noting that self-reported walking pace is considered a reliable indicator of gait speed and a well-established predictor of health outcomes.^40^ The generalizability of the observed interactions with other populations is limited due to the fact that the participants in the genetic analysis were only of European descent. Finally, due to the observational nature of this study, we cannot establish causality for the observed associations. A clinical trial for a walking pace intervention would be a valuable next step to determine causation and clinical relevance.

In this population-based study of adults in the UK, a faster walking pace is associated with lower risks of lung cancer occurrence and mortality. The study provides novel evidence that the association was modified by rs1815739 in *ACTN3* and rs7191721 in *RBFOX1*, underscoring the importance of considering genetic modifications in epidemiological studies.

## Data Availability

All data produced are available online at UK Biobank (https://www.ukbiobank.ac.uk/).

## Contributors

MYW: Conceptualization; Data curation; Formal analysis; Investigation; Methodology; Software; Visualization; Roles/Writing - original draft; Writing - review & editing.

JH, TW, PWCL: Conceptualization; Funding acquisition; Project administration; Software; Supervision; Writing - review & editing.

XQM, WT, RNS, HXP, SYW, JMR, and VN: Investigation; Methodology; Software; Supervision; Validation; Writing - review & editing.

All authors had full access to and verified all of the data. All authors approved the final version of the manuscript.

## Acknowledgements

This research was conducted using the UK Biobank resources under application 66137. We thank the participants for sharing their heath related information. Dr. Janice M. Ranson is supported by Alzheimer’s Research UK and the Alan Turing Institute/Engineering and Physical Sciences Research Council.

## Data Sharing Statement

The datasets are available to researchers through an open application via https://www.ukbiobank.ac.uk/register-apply/. The core scripts for data analyses and results visualization are posted at https://github.com/jielab/.

