## Supplementary figures and images for "Walking Pace, Sport Genes, and the Lung Cancer"

### Supplementary Figure 1

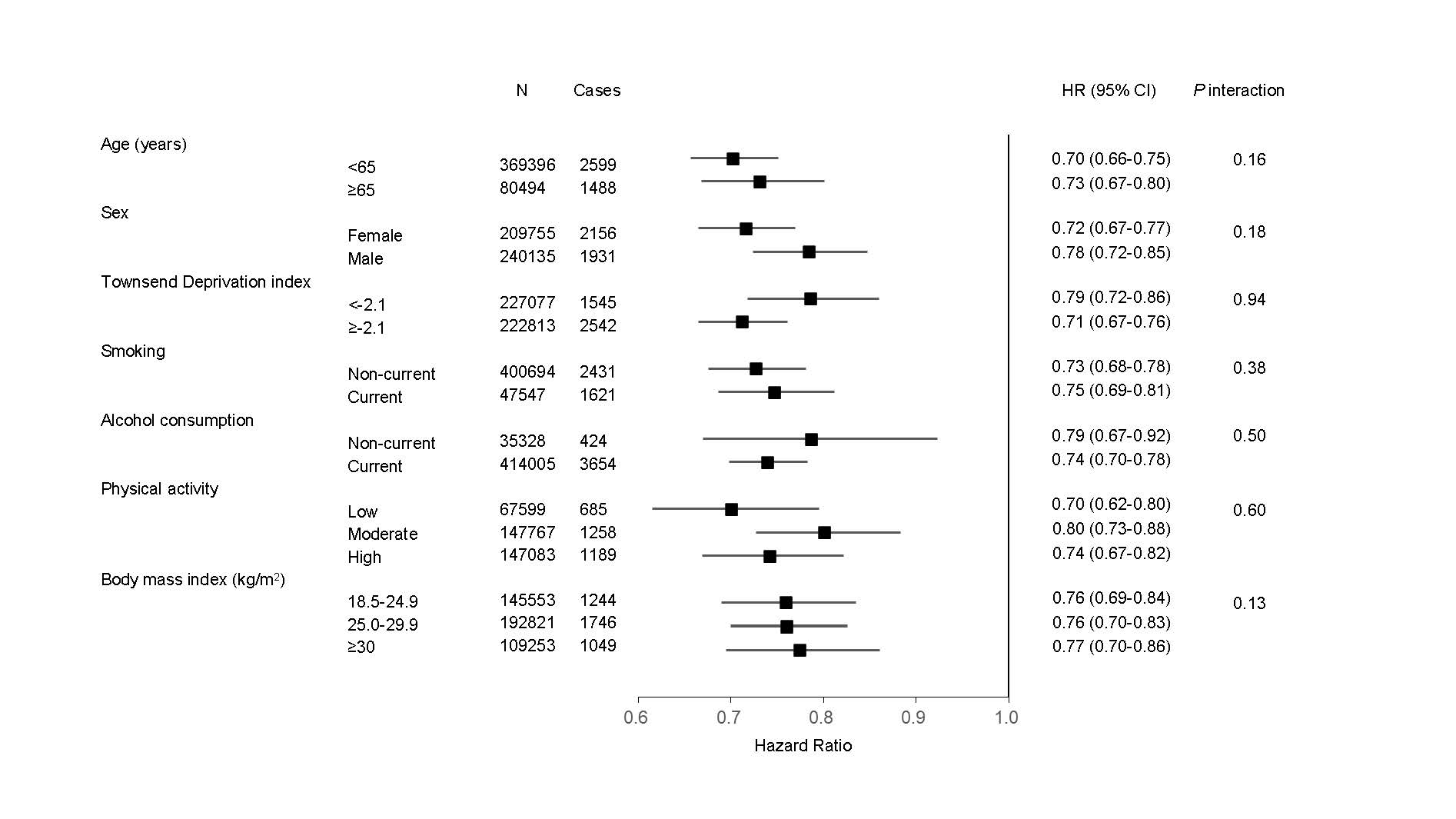

### Supplementary Figure 2

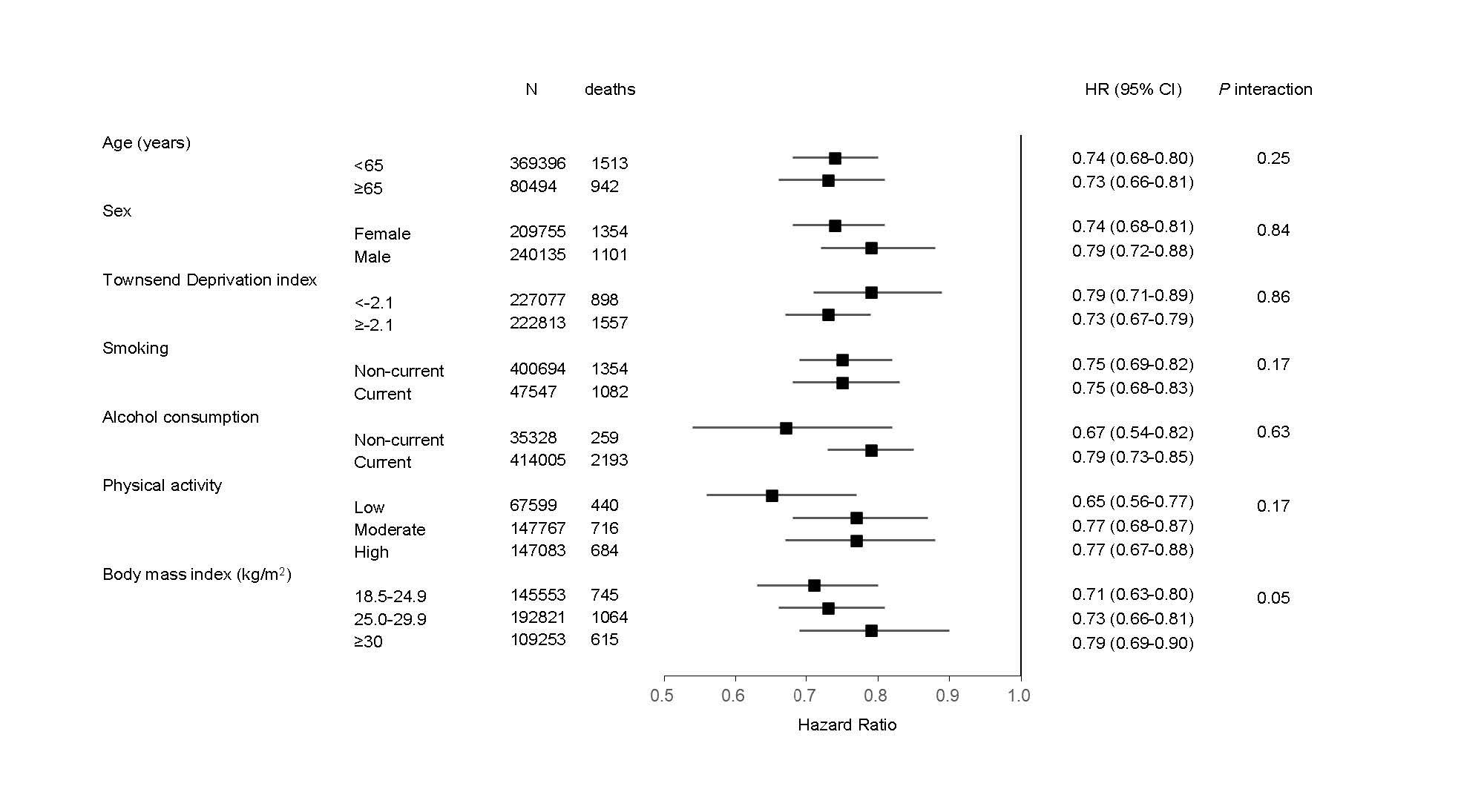

### Supplementary Figure 3

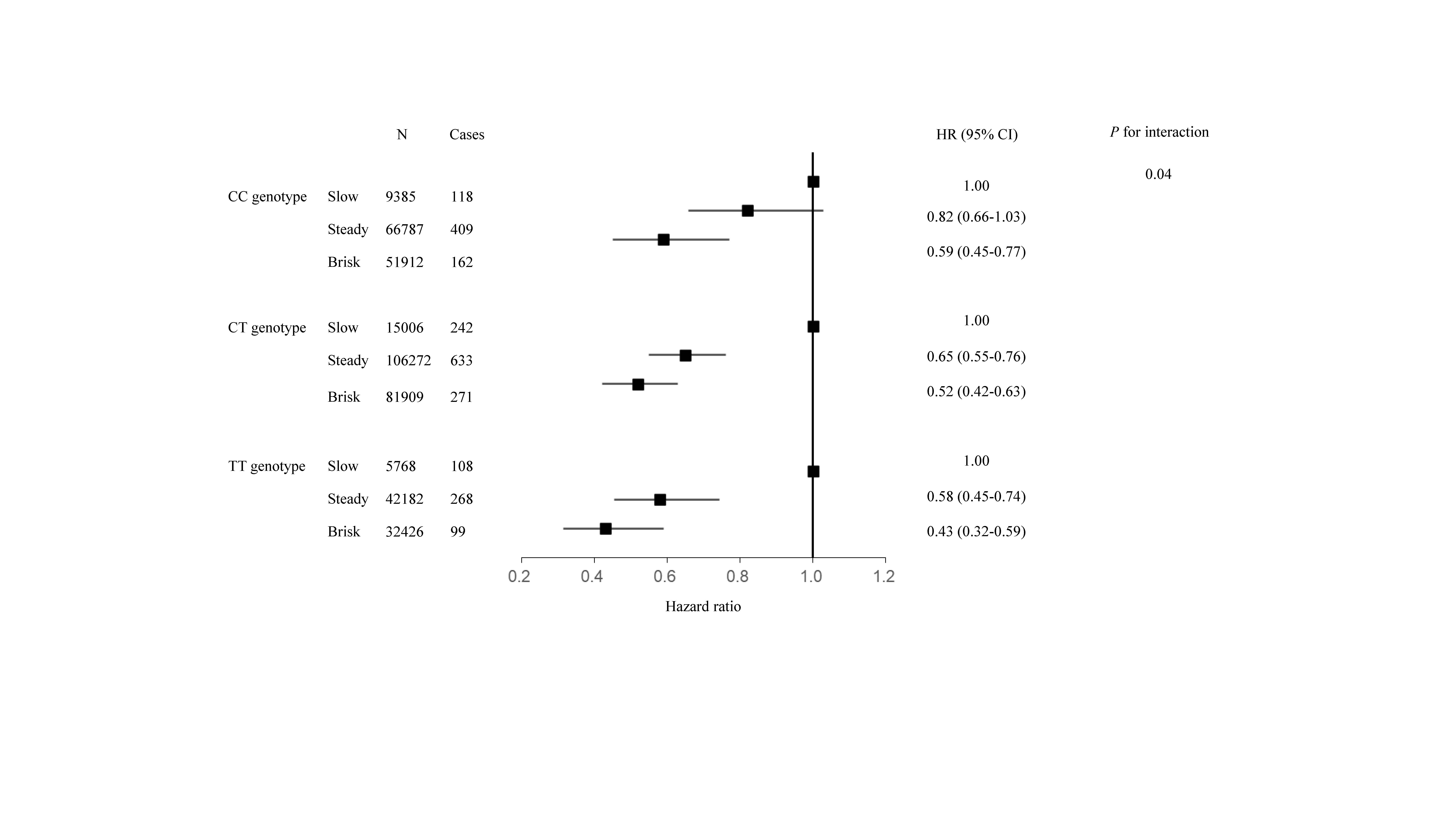

### Supplementary Figure 4

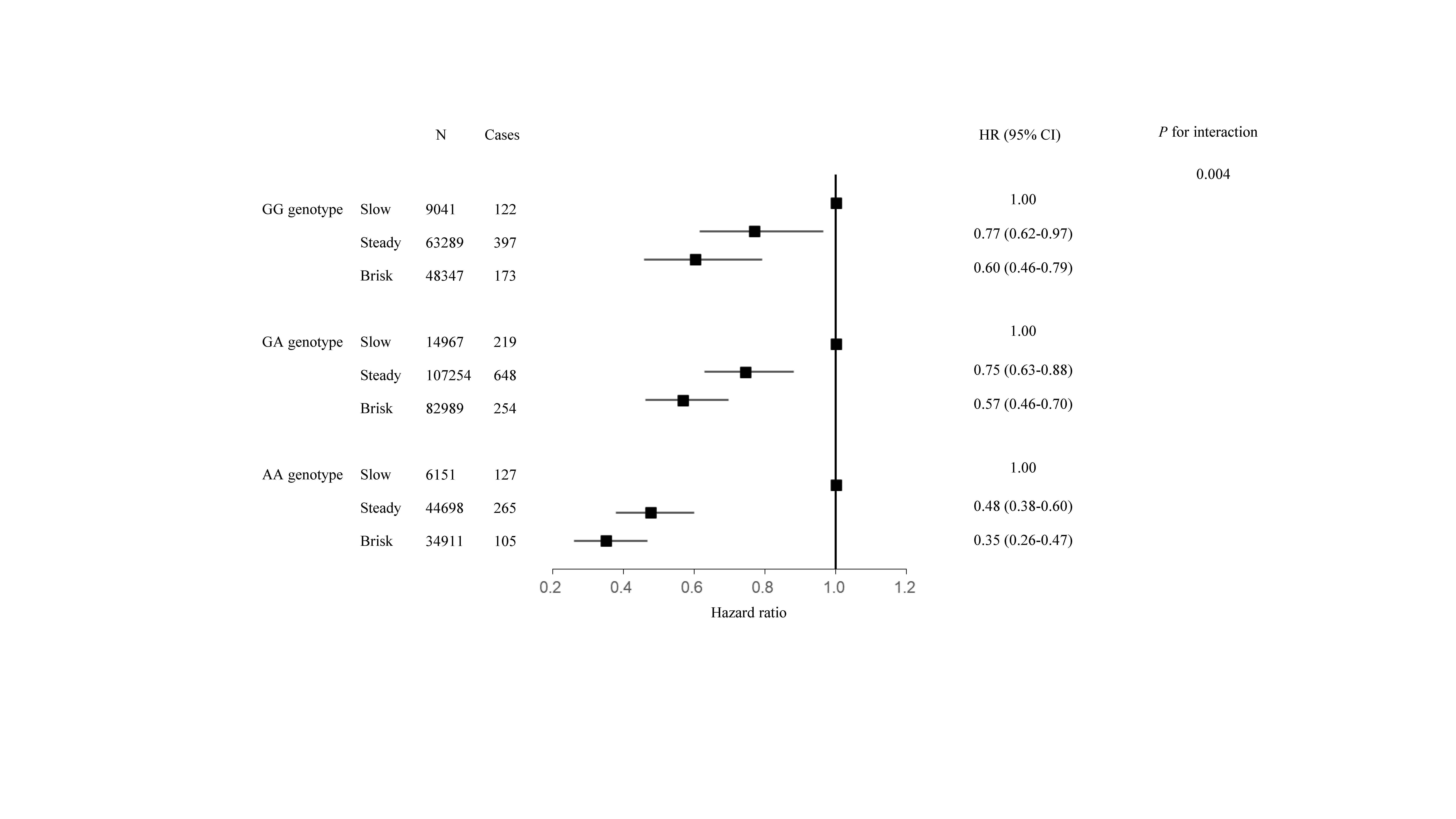
