## Supplementary Table 1-4 for "Walking Pace, Sport Genes, and the Lung Cancer"

Supplementary Table 1 Hazard ratio and 95% confidence interval of lung cancer risk for walking pace with further adjustment for particulate matter with diameters ≤2.5 µm

|  | Walking pace | | |  |
| --- | --- | --- | --- | --- |
|  | Slow | Steady | Brisk | *P* for trend |
| Lung cancer occurrence | 1.00 | 0.71 (0.65-0.78) | 0.55 (0.49-0.61) | <0.001 |
| Lung cancer mortality | 1.00 | 0.68 (0.61-0.76) | 0.52 (0.45-0.59) | <0.001 |

Supplementary Table 2 Hazard ratio and 95% confidence interval of lung cancer risk for walking pace with further adjustment for fat-free mass

|  | Walking pace | | |  |
| --- | --- | --- | --- | --- |
|  | Slow | Steady | Brisk | *P* for trend |
| Lung cancer occurrence | 1.00 | 0.71 (0.65-0.77) | 0.55 (0.50-0.62) | <0.001 |
| Lung cancer mortality | 1.00 | 0.68 (0.60-0.76) | 0.53 (0.46-0.60) | <0.001 |

Supplementary Table 3 Hazard ratio and 95% confidence interval of lung cancer risk for walking pace after excluding participants who had hypertension, diabetes, or cardiovascular disease at baseline

|  | Walking pace | | |  |
| --- | --- | --- | --- | --- |
|  | Slow | Steady | Brisk | *P* for trend |
| Lung cancer occurrence | 1.00 | 0.72 (0.59-0.87) | 0.56 (0.45-0.70) | <0.001 |
| Lung cancer mortality | 1.00 | 0.67 (0.52-0.85) | 0.53 (0.40-0.69) | <0.001 |

Supplementary Table 4 Hazard ratio and 95% confidence interval of lung cancer risk for walking pace after only including participants with a follow-up time of more than two years

|  | Walking pace | | |  |
| --- | --- | --- | --- | --- |
|  | Slow | Steady | Brisk | *P* for trend |
| Lung cancer occurrence | 1.00 | 0.73 (0.67-0.81) | 0.56 (0.50-0.63) | <0.001 |
| Lung cancer mortality | 1.00 | 0.69 (0.61-0.78) | 0.52 (0.45-0.61) | <0.001 |
